# Ocular Manifestations in a Cohort of Patients with Mpox in the Democratic Republic of the Congo 2007 - 2011

**DOI:** 10.1101/2025.04.07.25325069

**Authors:** Placide Mbala-Kingebeni, Nam V. Nguyen, Jean-Claude Mwanza, Jessica Carag, Susanne Linderman, Tolulope Fashina, Eddy Kinganda, Tony Wawina-Bokalanga, Amuri Aziza, Jean-Claude Makangara-Cigolo, Emmanuel Hasivirwe Vakaniaki, Yvon Anta, Jepsy Yango, Olivier Tshiani, Jason Kindrachuk, Laurens Liesenborghs, Martine Peeters, Eric Delaporte, Anne W. Rimoin, John W. Huggins, Therese Riu-Rovira, Steve M. Ahuka, James W. Martin, Ian Crozier, Steven Yeh, Jean-Jacques Muyembe

## Abstract

This report describes the ocular manifestations in a cohort of patients with Clade I mpox evaluated in the Democratic Republic of the Congo (DRC) from 2007 to 2011, as well as the clinical course of a pediatric patient with mpox associated with ocular disease. Among 216 patients, ocular manifestations were observed in 8.3% of the patients, including conjunctivitis, eyelid lesions, eyelid swelling, eye pain, orbital swelling, keratitis, and corneal staphyloma. These findings highlight the broad spectrum of ocular manifestations with potential sight-threatening consequences in patients with mpox.

## Background

Mpox, previously known as monkeypox, is a viral disease caused by the mpox virus (MPXV; also known as monkeypox virus), a double-stranded DNA virus classified within the *Orthopoxvirus* genus of the *Poxviridae* family.^1^ Thus far, two genetically distinct clades are known: Clade I and Clade II, with Clade I historically associated with higher morbidity and mortality than Clade II infection.^2^ Mpox has been known to be endemic to Central and Western Africa for decades, with Clade I infections predominantly reported in Central Africa and Clade II infections in Western Africa.^3^

Clinically, mpox is typically characterized by an initial prodromal phase that includes fever, lymphadenopathy, myalgia, fatigue, and headache, followed by the development of a characteristic vesiculopustular rash that begins on the face and spreads centrifugally.^3,4^ However, nonclassical mpox presentation associated with sexual/intimate contacts and sustained human transmission, including asynchronous lesion development and concentrated anogenital lesion localization, has been reported with increasing frequency.^5^ The global spread of Clade IIb mpox beginning in May 2022 led the World Health Organization (WHO) to declare a Public Health Emergency of International Concern (PHEIC) in July 2022.^6^ Although the PHEIC was lifted by May 2023, the epidemic of Clade IIb mpox continued globally, concomitant with the emergence of Clade Ib mpox, first detected in the Democratic Republic of the Congo (DRC) and subsequently spread to the neighboring countries and outside of Africa.^6^ The continuing spread of the epidemic prompted a renewed global PHEIC declaration in August 2024. Since 2022, there have been more than 120,000 confirmed mpox cases worldwide, and more than 70,000 suspected and probable mpox cases in the DRC alone since January 2024.^7^

Ocular manifestations are relatively frequent in patients with mpox, and they have been reported up to 23% in patients with Clade I mpox, predominantly affecting the ocular surface and anterior segment.^8^ Conjunctivitis has been the most commonly observed ocular manifestation, though severe ocular complications such as corneal ulceration, corneal perforation, or episcleritis may also occur, leading to significant visual impairment.^9-11^ The reported prevalence of ocular manifestations varies between 0.3% and 11% in Clade II mpox infections and 4.1% to 23.1% in Clade I mpox infections.^3^ Given the ongoing global mpox epidemic, which now includes Clade Ia, Ib, and IIb infections and the potential significant ocular morbidities associated with mpox, we aim to describe the ocular manifestations associated with Clade I mpox among patients admitted to the General Hospital of Kole, Sankuru in the DRC from 2007 to 2011, and we also detail the clinical course of a pediatric patient with mpox associated with ocular manifestations.

## Methods

The study was conducted in compliance with the Declaration of Helsinki, and informed consent was obtained from the patients participating in the study. The study was approved by the ethic committees of the Kinshasa School of Public Health in the DRC and the United States (US) Army Medical Research Institute of Infectious Diseases. This secondary analysis was performed to examine the spectrum of ocular manifestations in a cohort of patients with Clade I mpox in the DRC from 2007 to 2011.

The original study was a prospective observational study that was conducted at the General Hospital of Kole located in Sankuru District, Kasai Oriental Province, DRC, from 2007 to 2011. The details of the study design have been described previously.^4,12^ The region’s population primarily consists of hunters and farmers living in small villages within the tropical rainforest.^12^ Wild monkeys and rodents are often hunted by local communities with the risk of harboring viruses transmitted by direct contact with humors or carcasses of animals.^12^ Patients who met the WHO definition of mpox (suspected case, probable case, or confirmed case)^13^ and sought medical treatment at the hospital were invited to participate in the original study.^4,12^ Regardless of their enrollment status, all patients, who met the WHO definition of mpox, received the same medical management.^4,12^

A total of 216 patients met the inclusion criteria and were included in the original observational study.^4,12^ Collected demographic characteristics including age, and sex were analyzed for this secondary analysis. All enrolled patients underwent a comprehensive review of systems and physical examination at the time of evaluation for acute disease, and patients with ocular manifestations were reviewed and included in this secondary analysis. The clinical course of a pediatric patient with mpox associated with ocular manifestations was also reviewed and summarized herein. Statistical analyses were performed using Microsoft Excel (Microsoft Corp., Redmond, WA, USA). Categorical variables were presented as frequencies, while continuous variables were represented as median (IQR).

## Results

### Ocular manifestations in the study population

A total of 216 patients were included in the original study. The median age was 13 years, and 78 (36.1%) patients were female.^4^ Of 216 patients, 18 patients (8.3%) demonstrated ocular manifestations. The median age of those with ocular manifestations was 12.0 (4.6 – 21.0) years, with the majority (72.2%, n = 13) being pediatric patients (<18 years of age). Eight (44.4%) patients were female. Among the 18 patients with ocular manifestations, conjunctivitis was the most common finding (n=5, 27.8%), followed by eyelid lesions (n=4, 22.2%), eyelid swelling (n=3, 16.7%), eye pain (n=3, 16.7%), orbital swelling (n=1, 5.6%), keratitis (n=1, 5.6%), and corneal staphyloma (n=1, 5.6%). Among the patients with ocular symptoms, 3 pediatric patients (16.7%) also reported excessive tearing along with their ocular manifestations. One pediatric patient showed severe corneal opacity with diffuse facial scars from cutaneous lesions associated with mpox, and another pediatric patient demonstrated corneal staphyloma with corneal neovascularization after the acute stage of mpox. Available detailed information from one of the pediatric patients from the series is described below.

### Case report

A pediatric girl presented to the General Hospital of Kole with an acute onset of fever of 6 days duration and a vesicular rash 3 days prior. Past medical history included an unspecified diarrheal disease, and past surgical history was unremarkable. Dietary history was notable for Gambian rat consumption within 6 months of presentation. She had been vaccinated against measles but had never received vaccinations for smallpox or varicella zoster. Physical examination revealed eyelid edema with excessive tearing, cervical adenopathy, and numerous pustular lesions consistent with the diagnosis of mpox. Skin lesion count revealed a total of 141 lesions at various stages of healing with the majority of lesions localized to the head and facial regions. An external photograph of the right eye demonstrated significant eyelid swelling and conjunctival edema with deposits of whitish caseating material around the corneal limbus. The patient was admitted to the isolation unit for further management.

The patient underwent a comprehensive work-up including comprehensive metabolic panel (CMP), complete blood count (CBC) with differential, and urinalysis. In addition, a blood draw and oropharyngeal swab were collected for MPXV polymerase chain reaction (PCR) testing. CBC showed decreased anemia with normal white blood cell count, and CMP was unremarkable. PCR testing was positive for MPXV in both blood and oropharyngeal swab samples. The longitudinal PCR results from the blood draw and oropharyngeal swab samples are summarized in the **Figure 1**. Urinalysis revealed moderate protein and bilirubin with elevated urobilinogen.

**Figure 1:**
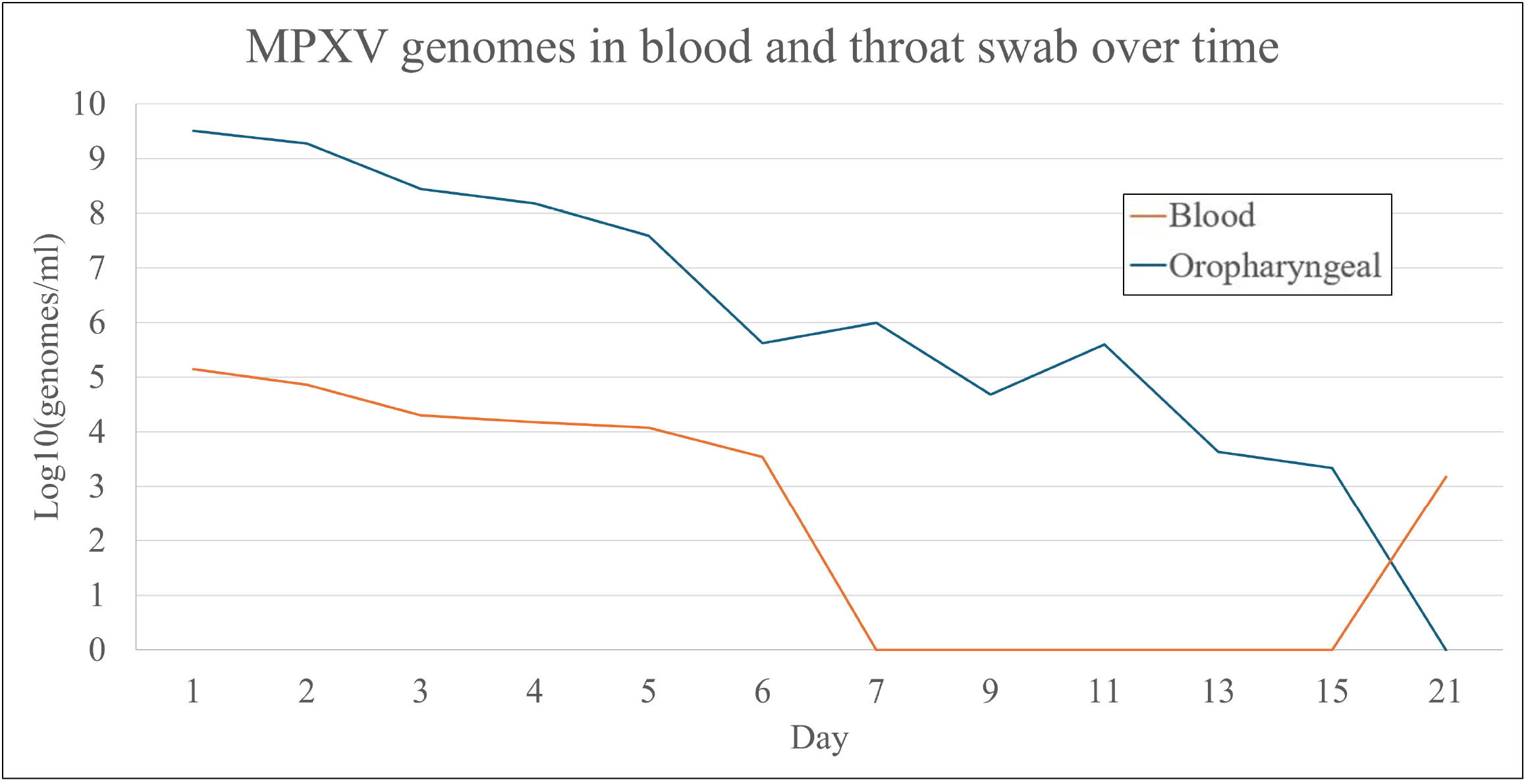
Monkeypox genomes concentration in blood and oropharyngeal swab over time in the patient described.

The patient was treated empirically for her ophthalmic findings with topical ophthalmic agents including chloramphenicol, tetracycline, ofloxacin, indomethacin, and atropine. Additionally, she also received prophylactic amoxicillin for secondary bacterial infections, and potassium permanganate for skin lesions. The eyelid edema resolved by Day 9 of the hospitalization; however, the patient demonstrated persistent narrowing of the palpebral fissure following the resolution of eyelid edema. By the time of discharge on Day 15, the majority of skin lesions resolved, and the patient was discharged in stable condition. The narrowing of the palpebral fissure was still present on Day 20 follow-up visit; however, it had resolved on Day 75 follow-up visit. She remained clinically stable on Day 75 follow-up visit.

## Discussion

This retrospective analysis of a landmark clinical characterization study in DRC (2007-2011) highlights the broad spectrum of ocular manifestations in patients with Clade I mpox, underscoring the potential for significant visual morbidity. Among 216 patients included in the original prospective study, we observed that 18 patients (8.3%) demonstrated ocular manifestations at the time of initial examination with conjunctivitis being the most common finding in this cohort, which is consistent with other reports in the literature.^8,9^ While follow-up for the patients with ocular manifestations in our study was not available, recent evidence from the literature has shown that keratitis may lead to significant visual morbidity from complications including corneal scarring, neovascularization, and perforation.^9-11^ Moreover, secondary ocular complications such as bacterial infections, or periorbital soft tissue infections can occur in the settings of keratitis and eyelid lesions, respectively. The patient described in this article demonstrated persistent narrowing of the palpebral fissure following the initial presentation with eyelid swelling and periorbital inflammation, illustrating the devastating effects of ocular manifestations in patients with mpox.

Notably, the majority of patients with ocular manifestations in our secondary analysis were pediatric patients with a median age of 5.7 years. Whether mpox may lead to a greater burden of ocular disease in pediatric patients warrants further studies, as other studies have reported a disproportionate number of pediatric patients with ocular manifestations compared to adult patients.^8,9^ Among 282 mpox patients described by *Jezek et*, more than 60% of patients was <15 years of age, and conjunctivitis was commonly reported, while keratitis was noted in 11 (4.4%) patients.^9^ Among these pediatric patients, one girl experienced bilateral blindness, three children reported unilateral blindness, and six other children reported various degrees of visual impairment due to corneal opacities.^9^ *Hughes et al* also reported 23.1% of patients with conjunctivitis in a cohort of 68 patients with 61.8% of patients less than 10 years old. Notably, the patients described in these two studies were affected by the Clade I variant, which tends to be associated with a higher morbidity than the Clade II variant.^3,8,9^ With the ongoing circulation of Clade I MPXV, and given the risks of visual impairment in children with mpox, health care providers should be aware of ocular complications when evaluating children with mpox. Prompt referrals to an ophthalmologist should be made to prevent visual loss in pediatric patients, who are also at a high risk of developing amblyopia.

The pathogenesis of ocular manifestations in patients with mpox remains poorly understood; however, recent evidence from cases of mpox-associated ocular disease has shown signs of persistence of MPXV genomes within extra- and intraocular ocular fluid that could play a role in disease pathogenesis.^11,14^ Other studies reported the persistence of MPXV genomes within the ocular surface and aqueous humor samples at 145 days and 8 months, respectively, after the acute infection.^14,15^ Further studies are needed to explore the role of MPXV genomes’ persistence in extra- and intraocular fluid and their role in the pathogenesis of ocular manifestations in patients with mpox.

This study is limited by its retrospective nature, lack of visual acuity assessment and slit lamp biomicroscopic evaluation, and the resource constraints of the study setting. The remote location of this study with limited ophthalmic services made the detailed assessment difficult. Nonetheless, external photographic documentation was able to capture severe ophthalmic disease likely associated with vision impairment in patients with mpox. As Clade I mpox was circulating in Kole, DRC at the time of this report, further study of both Clade Ia and Ib mpox in the ongoing outbreak is particularly relevant for vision health and quality-of-life for patients with mpox among patients under 18 years old as they might be at higher risk. The broad spectrum of ocular manifestations including conjunctivitis, keratitis, and periorbital manifestations in patients with mpox highlights the breadth of ophthalmic subspecialty care and management required.

These resource requirements may be particularly challenging within resource-limited settings where mpox transmission has been documented. Heightened surveillance measures with further research for ocular manifestations associated with mpox remain critical given the expanding number of cases in the DRC, neighboring countries in sub-Saharan Africa, and countries previously unaffected by mpox that have recently reported Clade I mpox.

## Data Availability

All data produced in the present study are available upon reasonable request to the authors

## Acknowledgments

Not applicable.

## Data sharing statement

Data requests can be made to the corresponding author with a supplemental proposal outlining the proposed analysis. If permission is granted, deidentified study participant data in an Excel format will be made available. Data requests will be reviewed pending publication.

## Ethical approval

This article was conducted in accordance with the Declaration of Helsinki. The collection and evaluation of all protected patient health information was performed in a Health Insurance Portability and Accountability Act (HIPAA)-compliant manner.

## REFERENCES

1. Gessain A, Nakoune E, Yazdanpanah Y. Monkeypox. N Engl J Med. Nov 10 2022;387(19):1783–1793. doi:10.1056/NEJMra2208860

2. Okwor T, Mbala PK, Evans DH, Kindrachuk J. A contemporary review of clade-specific virological differences in monkeypox viruses. Clin Microbiol Infect. Dec 2023;29(12):1502–1507. doi:10.1016/j.cmi.2023.07.011

3. Begley J, Kaftan T, Song H, et al. Ocular Complications of Mpox: Evolving Understanding and Future Directions. Int Ophthalmol Clin. Oct 1 2024;64(4):15–22. doi:10.1097/IIO.0000000000000536

4. Pittman PR, Martin JW, Kingebeni PM, et al. Clinical characterization and placental pathology of mpox infection in hospitalized patients in the Democratic Republic of the Congo. PLoS Negl Trop Dis. Apr 2023;17(4):e0010384. doi:10.1371/journal.pntd.0010384

5. Van Dijck C, Hoff NA, Mbala-Kingebeni P, et al. Emergence of mpox in the post-smallpox era-a narrative review on mpox epidemiology. Clin Microbiol Infect. Dec 2023;29(12):1487–1492. doi:10.1016/j.cmi.2023.08.008

6. WHO. WHO Director-General declares mpox outbreak a public health emergency of international concern. World Healt Organization (WHO). Accessed 01/18/2025, 2025. https://www.who.int/news/item/14-08-2024-who-director-general-declares-mpox-outbreak-a-public-health-emergency-of-international-concern

7. WHO. Global Mpox Trends. Accessed 03/25/2025, 2025. https://worldhealthorg.shinyapps.io/mpx_global/

8. C Hughes AM, E Pukuta, S Karhemere, B Nguete, R Shongo Lushima, J Kabamba, M Balilo, JJ Muyembe Tamfum, O Wemakoy, J Malekani, B Montoe, IK Damon, MG Reynolds Ocular complications associated with acute monkeypox virus infection, DRC. International Journal of Infectious Diseases 04/2014 2014;21:276–277.

9. Jezek Z, Szczeniowski M, Paluku KM, Mutombo M. Human monkeypox: clinical features of 282 patients. J Infect Dis. Aug 1987;156(2):293–8. doi:10.1093/infdis/156.2.293

10. Carrubba S, Geevarghese A, Solli E, et al. Novel severe oculocutaneous manifestations of human monkeypox virus infection and their historical analogues. Lancet Infect Dis. May 2023;23(5):e190–e197. doi:10.1016/S1473-3099(22)00869-6

11. Nguyen MT, Mentreddy A, Schallhorn J, et al. Isolated Ocular Mpox without Skin Lesions, United States. Emerg Infect Dis. Jun 2023;29(6):1285–1288. doi:10.3201/eid2906.230032

12. Mbala PK, Huggins JW, Riu-Rovira T, et al. Maternal and Fetal Outcomes Among Pregnant Women With Human Monkeypox Infection in the Democratic Republic of Congo. J Infect Dis. Oct 17 2017;216(7):824–828. doi:10.1093/infdis/jix260

13. WHO. Mpox (Monkeypox) outbreak toolbox. WHO. Accessed 02/13/2025, 2025. https://www.who.int/emergencies/outbreak-toolkit/disease-outbreak-toolboxes/mpox-outbreak-toolbox

14. Raccagni AR, Clemente T, Ranzenigo M, Cicinelli MV, Castagna A, Nozza S. Persistent ocular mpox infection in an immunocompetent individual. Lancet Infect Dis. Jun 2023;23(6):652–653. doi:10.1016/S1473-3099(23)00266-9

15. Finamor LPS, Mendes-Correa MC, Rinkevicius M, et al. Ocular manifestations of Monkeypox virus (MPXV) infection with viral persistence in ocular samples: A case series. Int J Infect Dis. Sep 2024;146:107071. doi:10.1016/j.ijid.2024.107071

